# Complement 3a Receptor 1 on Macrophages and Kupffer cells is not required for the Pathogenesis of Metabolic Dysfunction-Associated Steatotic Liver Disease

**DOI:** 10.1101/2024.06.26.24309550

**Authors:** Edwin A. Homan, Ankit Gilani, Alfonso Rubio-Navarro, Maya A. Johnson, Odin M. Schaepkens, Eric Cortada, Renan Pereira de Lima, Lisa Stoll, James C. Lo

## Abstract

Together with obesity and type 2 diabetes, metabolic dysfunction-associated steatotic liver disease (MASLD) is a growing global epidemic. Activation of the complement system and infiltration of macrophages has been linked to progression of metabolic liver disease. The role of complement receptors in macrophage activation and recruitment in MASLD remains poorly understood. In human and mouse, *C3AR1* in the liver is expressed primarily in Kupffer cells, but is downregulated in humans with MASLD compared to obese controls. To test the role of complement 3a receptor (C3aR1) on macrophages and liver resident macrophages in MASLD, we generated mice deficient in C3aR1 on all macrophages (C3aR1-MjKO) or specifically in liver Kupffer cells (C3aR1-KpKO) and subjected them to a model of metabolic steatotic liver disease. We show that macrophages account for the vast majority of *C3ar1* expression in the liver. Overall, C3aR1-MjKO and C3aR1-KpKO mice have similar body weight gain without significant alterations in glucose homeostasis, hepatic steatosis and fibrosis, compared to controls on a MASLD-inducing diet. This study demonstrates that C3aR1 deletion in macrophages or Kupffer cells, the predominant liver cell type expressing *C3aR1*, has no significant effect on liver steatosis, inflammation or fibrosis in a dietary MASLD model.

## Introduction

Obesity and related metabolic diseases such as type 2 diabetes (T2D) and metabolic dysfunction-associated steatotic liver disease (MASLD) remain a worldwide epidemic with increasing prevalence^1,2^. MASLD describes the constellation of hepatic lipid deposition, inflammation, and fibrosis associated with obesity and T2D that ultimately leads to MASH cirrhosis, which has become the leading cause of liver transplantation in the United States^3–6^. Notably, MASLD is increasingly recognized as an important risk-enhancing factor for atherosclerotic cardiovascular disease^7,8^.

Liver macrophages help to maintain hepatic homeostasis and consist of embryo-derived resident macrophages called Kupffer cells, which self-renew and do not migrate, or peripheral monocyte-derived macrophages, which infiltrate into liver tissue upon metabolic or toxic liver injury and under certain circumstances can take on Kupffer cell-like identity^9–13^. In obesity, bone marrow-derived myeloid cells migrate to the steatotic liver, and pro-inflammatory recruited macrophages are postulated to drive the progression of MASLD to MASH^14^. Spatial proteogenomics reveals a population of lipid-associated macrophages near bile canaliculi that is induced by local lipid exposure and drives fibrosis in steatotic regions of murine and human liver^15^. In addition, deep transcriptomic profiling in human MASLD has identified candidate gene signatures for steatohepatitis and fibrosis with possible therapeutic implications^16^.

Activation of the body’s complement system leads to increased cell lysis, phagocytosis, and inflammation^17^, and it is increasingly recognized as an important contributor to regulation of metabolic disorders such as T2D and MASLD^18,19^. In human liver biopsies, higher lobular inflammation scores correlate with activation of the complement alternative pathway^20^, which can signal *via* the C3a receptor 1 (C3aR1), a G_i_-coupled G protein-coupled receptor^21^. The complement 3 polypeptide (C3) is cleaved by C3 convertase to the activated fragment, C3a, which then binds C3aR1^22^. Complement factor D (CFD), also known as the adipokine adipsin, is the rate-limiting step in the alternative pathway of complement activation^23,24^.

Several studies have reported opposing roles of adipsin and C3aR1 on hepatic steatosis in diet-induced obesity^25–27^. Our lab has found that adipsin/CFD is critical for maintaining pancreatic beta cell mass and function^28,29^. Murine obese and diabetic models such as *db/db* mice and high fat diet (HFD) feeding result in very low circulating adipsin^23^. Replenishing adipsin in *db/db* mice raises levels of C3a and insulin, lowers blood glucose levels, and inhibits hepatic gluconeogenesis^28^. However, whole-body deletion of C3aR1 decreases macrophage infiltration and activation in adipose tissue, protects from HFD-induced obesity and glucose intolerance, and decreases hepatic steatosis and inflammation^30^. In a model of fibrosing steatohepatitis, bone marrow-derived macrophages were found to activate hepatic stellate cells, which was blunted in whole-body C3aR1 KO mice^31^.

In the present study we aim to explore the macrophage-specific effect of complement receptor signaling in MASLD pathogenesis. To determine the consequences of macrophage and Kupffer cell ablation of C3aR1, we use a murine dietary model of MALFD/MASH, the Gubra Amylin Nash (GAN) diet, which has macronutrient similarities to the Western diet and produces similar histologic and transcriptomic changes to human MASLD/MASH^32–34^.

## Results

### C3AR1 is expressed in human and mouse liver, primarily in Kupffer cells

In the scRNA-Seq database, Human Protein Atlas, *C3AR1* is broadly expressed throughout the body, with increased abundance in tissues rich in immunologic cell types, such as bone marrow and appendix (Fig. 1A)^35^. In a single-cell transcriptomic database of healthy human liver, *C3AR1* expression predominates in the macrophage and Kupffer cell population, with minimal-to-undetectable *C3AR1* expression in hepatocytes or hepatic stellate cells by scRNA-Seq (Fig. 1B)^36^. In the mouse liver scRNA-Seq database, Tabula Muris, *C3ar1* is similarly expressed primarily in Kupffer cells (Fig. S1)^37^.

**Figure 1.**
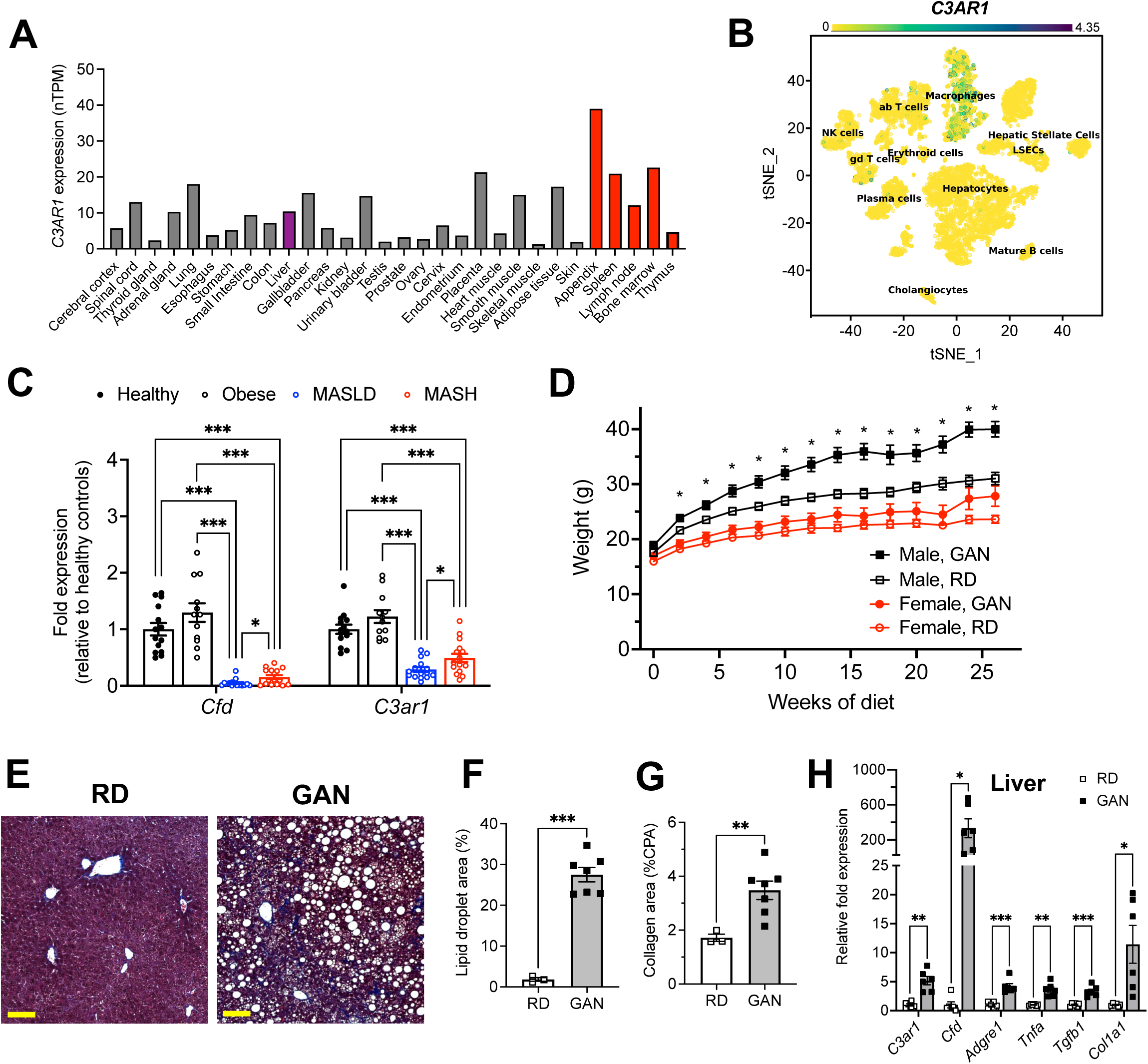
C3AR1 is found in macrophages, is modulated by MASLD/MASH in humans, and is induced by a murine dietary model of MASH. A) Relative *C3AR1* human tissue expression level by tissue, derived from deep sequencing of the mRNA combined dataset (HPA and GTEx) in the Human Protein Atlas, shown as normalized transcripts per million (nTPM). Liver is highlighted in purple and immunologic tissues are highlighted in red. B) Single-cell RNA sequencing distribution of *C3AR1* expression in human liver (tSNE, t-distributed Stochastic Neighbor Embedding). C) Analysis of *CFD* and *C3AR1* expression from liver biopsy samples in patients with MASH, MASLD, obesity without MASLD, and age-matched healthy controls (n = 12-16 per group, Welch *t* test with Holm-Šídák correction for multiple comparisons). D) Weight curve in male and female flox/flox control mice placed on GAN high-fat diet compared to regular diet (RD) controls (males, n = 7; females, n = 6). E) Representative liver section staining by Masson’s Trichrome in male control mice on RD or GAN diet for 28 weeks (scale bar = 100 mm). F) Lipid droplet area quantification in liver sections from male control mice, excluding vessel lumens (RD, n = 3; GAN, n = 7). G) Collagen area quantification in liver sections of male control mice (RD, n = 3; GAN, n = 7). H) Gene expression of key macrophage or fibrosis genes in male control mice on GAN or RD (n = 6 per group). Unpaired two-tailed Student’s *t* test (Except 1C as above). Annotations: *, p < 0.05; **, p < 0.01; ***, p < 0.001

### Hepatic CFD and C3AR1 are downregulated in human MASLD/MASH

We also examined data from Suppli and coworkers, who performed bulk transcriptomic analysis of human liver samples from an age-matched cohort of healthy controls and obese controls without MASLD, as well as MASLD and MASH patients without cirrhosis^38^. Both *CFD and C3AR1* were unchanged in obese subjects without MASLD compared to healthy controls, but both *CFD* and *C3AR1* were significantly downregulated in liver biopsies from both MASLD and MASH patients compared to both healthy controls and obese subjects without MASLD (Fig. 1C). Interestingly, both *CFD* and *C3AR1* levels were slightly higher in MASH individuals compared to those with MASLD only.

### Murine MASH model recapitulates key features of human MASH

At 5 weeks of age, we subjected *C3ar1* flox/flox control mice to standard regular diet (RD) or GAN diet^32,33^. After 28 weeks of GAN diet, male mice gained body weight compared to RD (Fig. 1D), primarily as fat mass (Fig. S2-3), but weight gain in female GAN-fed mice was attenuated. Histologic signs of MASLD were present in GAN-fed mice (Fig. 1E), most notably hepatic steatosis and hepatocyte ballooning (Fig. 1F), and liver fibrosis measured by collagen deposition nearly doubled with GAN compared to RD (Fig. 1G). Both hepatic *C3ar1* and *Cfd* gene expression were robustly increased on GAN compared to RD, as were markers of macrophage infiltration, hepatic inflammation, and fibrosis, including collagen gene expression, indicating progression to fibrotic MASH (Fig. 1H). In female control mice on GAN diet, there were no significant differences in *C3ar1* expression or other gene markers, though there was a nonsignificant trend toward increased inflammation and fibrosis compared to regular diet (Fig. S4).

### Macrophage-specific C3aR1 deletion does not alter glucose homeostasis

Owing to higher levels of *C3ar1* in murine MASLD and the differential regulation of *C3AR1* gene in MASLD humans, this motivated us to interrogate the role of pathophysiological role of *C3ar1* in macrophages in MASLD. We generated transgenic mice with macrophage-specific deletion of C3aR1 by crossing *C3ar1* floxed mice with LysM-Cre transgenic mice (C3aR1-MjKO) to target both liver resident macrophages and recruited monocytes. *C3ar1* floxed mice were used as controls. Successful deletion of *C3ar1* in macrophages from the C3aR1-MjKO mouse was confirmed by quantitative RT-PCR of isolated peritoneal macrophages that were F4/80+ and CD68+ by fluorescence-activated cell sorting (Fig. 2A). In liver tissue, *C3ar1* expression was reduced by ∼88% in both male and female C3aR1-MjKO (Fig. 2B). These results indicate that macrophages account for the vast majority of *C3ar1* expression in the liver.

**Figure 2.**
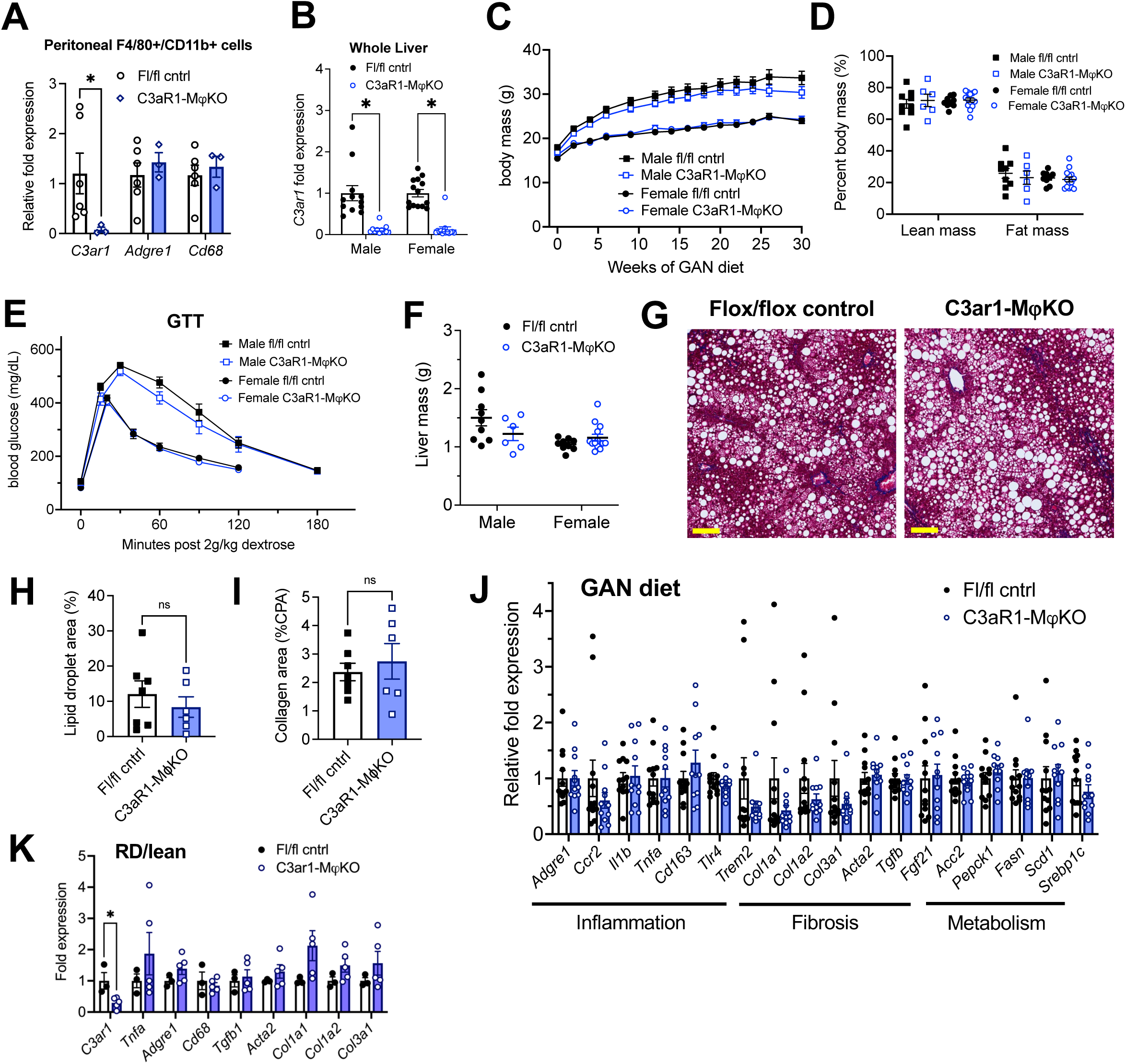
C3aR1 deletion in all macrophages does not affect weight gain, glucose homeostasis, liver steatosis or fibrosis. A) Expression of *C3ar1* in isolated peritoneal F4/80+/CD68+ cells from flox/flox control mice (n = 6) or C3aR1-MjKO male mice (n = 3). B) Expression of *C3ar1* in whole liver from control or C3aR1-MjKO mice (n = 11-12 per male group, n = 13-14 per female group). C) Body mass curve of control or C3aR1-MjKO mice on GAN high-fat diet starting at 5 weeks of age (n = 11-12 per male group, n = 14 per female group). D) Body composition analysis by EchoMRI in control or C3aR1-MjKO mice after 30 weeks GAN diet (n = 6-9 per male group, n = 9-13 per female group). E) Glucose tolerance test in control or C3aR1-MjKO mice with 14h fast after 28 weeks GAN diet (n = 6-9 per male group, n = 9-14 per female group). F) Liver mass in control or C3ar1-MjKO male mice at time of euthanasia after 30 weeks GAN diet (n = 6-9 per male group, n = 9-14 per female group). G) Representative liver section staining by Masson’s Trichrome in male control or C3ar1-MjKO mice (scale bar = 100 mm). H) Lipid droplet area in liver sections from male control or C3ar1-MjKO mice, excluding vessel lumens (n = 6-7 per group). I) Collagen area in liver sections from male control or C3ar1-MjKO mice (n = 6-7 per group). J,K) Relative mRNA expression of key markers for inflammation, fibrosis, and liver metabolism in liver from male control or C3ar1-MjKO mice after 30 weeks of either GAN (J) diet (n = 11-12 per group) or regular (K) diet (n = 3-5 per group). Unpaired two-tailed Student’s *t* test: Student’s *t* test: *, p < 0.05.

When placed on GAN diet, there was no significant difference in weight gain between control and C3aR1-MjKO mice (Fig. 2C). There was similarly no difference in percent lean or fat mass between these mice (Fig. 2D). Glucose tolerance tests performed in fasted mice after 27 weeks GAN diet found no significant differences between control and C3aR1-MjKO mice (Fig. 2E). There was also no difference in insulin sensitivity as measured by insulin tolerance tests in male mice (Fig. S5). Insulin resistance as measured by comparing the ratio of fasting glucose level to fasting insulin level (HOMA-IR) was also unchanged between controls and C3aR1-MjKO mice (Fig. S6). Circulating serum ALT levels were unchanged in male control and C3aR1-MjKO mice on GAN diet (Fig. S7).

### Macrophage-specific C3aR1 deletion does not significantly impact hepatic steatosis or fibrosis

Liver samples collected after 28-30 weeks of GAN or regular diet did not show significant differences in liver mass between control and C3aR1-MjKO mice (Fig. 2F). Male mice on GAN diet developed similar qualitative appearance on histology (Fig. 2G), and slide image analysis showed similar proportions of lipid droplet area and collagen area (Figs. 2H, 2I). This indicates that there were no significant differences in steatosis or fibrosis between GAN-fed control and C3aR1-MjKO male mice. While *C3ar1* expression was markedly reduced in the C3aR1-MjKO liver tissue (Fig. 2B), there were no detectable gene expression changes in markers of fibrosis, inflammation, or lipid handling on either GAN or regular diet (Fig. 2J-K). Similarly, in female mice there were also no significant differences between control and C3aR1-MjKO mouse liver on either GAN or regular diet in a subset of key gene markers of fibrosis or inflammation (Fig. S8).

### Kupffer cell-specific C3aR1 deletion does not alter weight gain or glucose homeostasis

To explore whether there may be competing effects between recruited monocytes and liver resident macrophages (Kupffer cells), we next generated Kupffer cell-specific C3aR1 knockout mice (C3aR1-KpKO) by crossing *C3ar1* floxed mice to Clec4f-Cre transgenic mice and fed them GAN diet. *C3ar1* floxed mice were used as controls. Body weight gain was similar between genotypes for both male and female mice (Fig. 3A), and there was no difference in body composition between control and C3aR1-KpKO mice on GAN diet (Fig. 3B). There was similarly no significant difference in glucose homeostasis between the genotypes during a glucose tolerance test (Fig. 3C).

**Figure 3.**
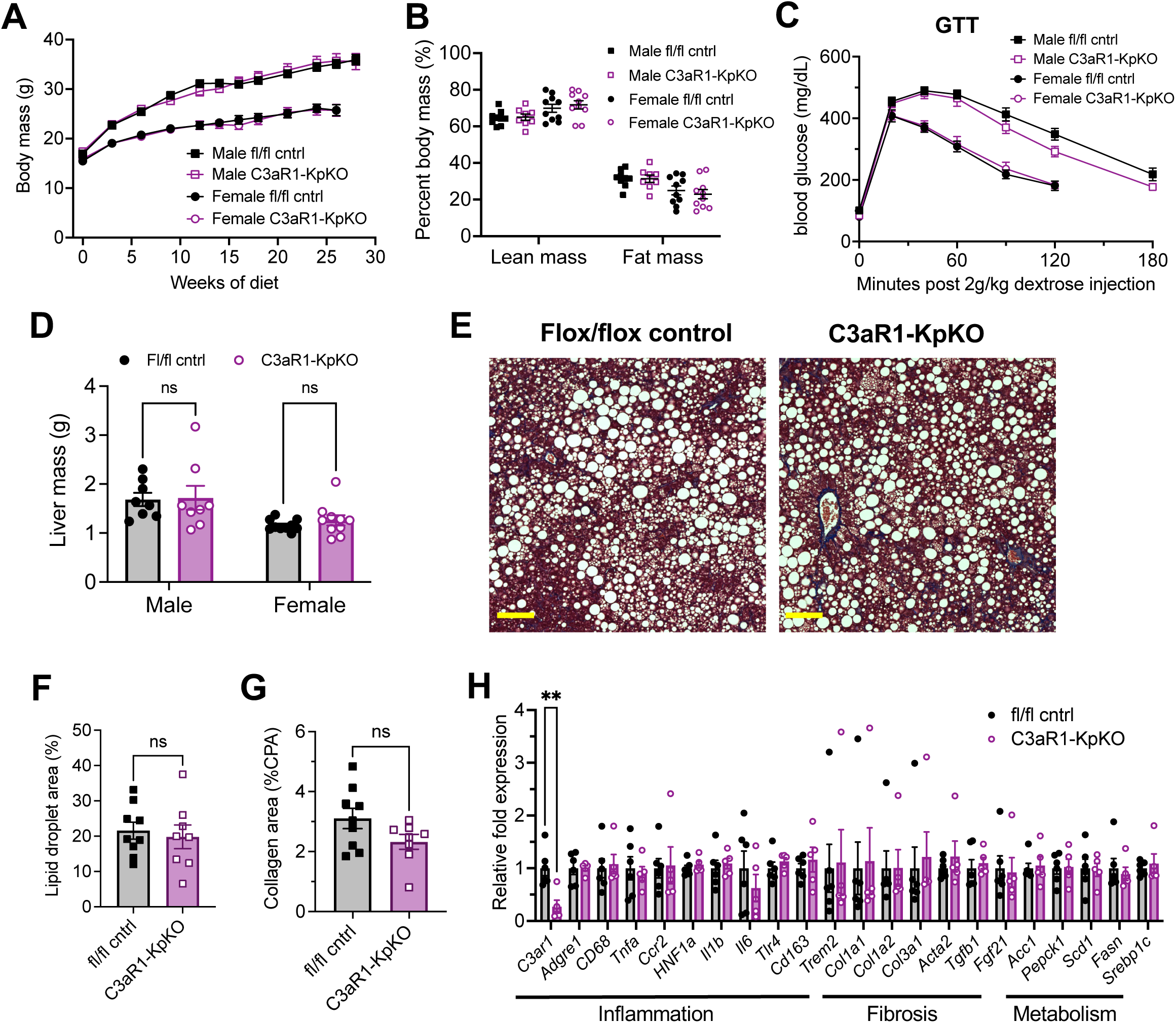
C3aR1 deletion in Kupffer cells does not affect weight gain, glucose homeostasis, liver steatosis or fibrosis. A) Body mass curve on GAN diet in flox/flox control or C3aR1-KpKO mice beginning at 5 weeks of age (n = 8-10 per group). B) Body composition analysis by EchoMRI in control or C3aR1-KpKO mice after 28 weeks GAN diet (n = 8-10). C) Glucose tolerance test in control or C3aR1-KpKO mice with 14h fast after 26 weeks GAN diet (n = 8-10). D) Liver mass in control or C3aR1-KpKO male mice at time of euthanasia after 30 weeks GAN diet (n = 8-10). E) Representative liver section staining by Masson’s Trichrome in control or C3aR1-KpKO male mice (scale bar = 100 mm). F) Lipid droplet area quantified on liver sections of control or C3aR1-KpKO male mice, excluding vessel lumens (n = 8-9). G) Collagen area quantified on whole liver section of control or C3aR1-KpKO male mice (n= 8-9). H) Relative gene expression in male control or C3aR1-KpKO mice after 30 weeks GAN diet (n = 5-6). Unpaired two-tailed Student’s *t* test: **, p < 0.01.

### Kupffer cell-specific C3aR1 deletion does not significantly impact hepatic steatosis or fibrosis

Liver mass was not significantly different between control and C3aR1-KpKO mice on GAN diet (Fig. 3D). Liver sections appeared qualitatively similar by histology stained with Masson’s trichrome (Fig. 3E). There were similar levels of hepatic steatosis in these mice as measured by percent lipid droplet area (Fig. 3F). When measured by collagen proportional area, there was no significant differences in liver fibrosis between C3aR1-KpKO and control mice (Fig. 3G). While *C3ar1* expression was reduced by 73% in liver tissue of C3aR1-KpKO mice, there were no significant differences in expression of inflammatory, fibrotic, or lipid handling gene markers (Fig. 3H). *C3ar1* expression similarly decreased by ∼90% in liver tissue of female C3aR1-KpKO mice fed regular diet compared to control mice (Fig. S9). These data also indicate that Kupffer cells account for ∼80% of hepatic *C3ar1* gene expression in our mouse model of MASLD/MASH.

## Discussion

Overall, we found that macrophage or Kupffer cell expression of *C3ar1* does not impact body weight gain or histologic/transcriptomic features of MASLD/MASH in a murine dietary model. Deletion of C3aR1 in the macrophage population throughout the body, or specifically in Kupffer cells, did not affect weight gain, glucose homeostasis, or extent of hepatic steatosis/fibrosis. With long term GAN diet feeding that has been previously shown to model human MASLD/MASH, we did not observe significant differences in liver abnormalities with the KO mice.

Our findings in macrophage-specific C3aR1 KO mice contrast with prior observations in whole-body C3aR1 KO mice^30^, which are protected from diet-induced obesity, have improved glucose tolerance, and exhibit decreased hepatic steatosis. In both our macrophage-and Kupffer cell-specific C3aR1 KO mice, which had similar degrees of obesity compared to controls, there was no detectable effect on liver steatosis or fibrosis despite the near abrogation of *C3ar1* expression. This raises the possibility that the lower levels of hepatic steatosis and insulin resistance previously observed in the whole body C3aR1 KO mice may be secondary to protection from obesity. Protection from diet-induced obesity in whole-body C3aR1 KO mice may be mediated by a non-macrophage cell type, since our macrophage-specific C3aR1 KO mice were not afforded this protection. The *C3ar1*-expressing cell types that promote obesity and MASLD remains to be determined.

Our laboratory recently reported sex-dependent regulation of thermogenic adipose tissue mediated by adipocyte-derived C3aR1^39^. However, no such sexual dimorphism was observed in hepatic expression of key MASH genes in response to GAN diet in our macrophage-or Kupffer cell-specific C3aR1-deficient mice. Other work has suggested possible compensatory effects from its sister anaphylatoxin receptor C5aR1, with increased cold-induced adipocyte browning and attenuated diet-induced obesity seen in C3aR1/C5aR1 double KO mice^40^.

The strengths of our study include careful metabolic and transcriptomic phenotyping of cell type-specific transgenic mice. Some limitations were our use of a single MASLD dietary model and our focus on the C3aR1 pathway. While the GAN diet recapitulates many features of human MASH due to its similarity to Western diet^34^, relatively low levels of fibrosis were seen in our study, potentially related to initiating the diet at young age; more rapid fibrosis induction has been seen when GAN diet is initiated at older ages^41^. It is possible that in other models of liver injury that we did not test (e.g., short-term treatment with a hepatotoxin such as carbon tetrachloride),^42^ there may be differences in liver injury in mice lacking *C3ar1* in macrophages. However, the GAN diet model has been shown to better parallel the gene expression changes in human MAFLD/MASH.^33^ Lastly, while *C3AR1*/*C3ar1* expression is very low in non-macrophage cells (Fig. B, S1), C3aR1 signaling on other hepatic cell types not explored in this study, such as hepatic stellate cells, could mediate the observed effect in the whole-body C3aR1 KO mouse.

Deletion of C3aR1 in macrophages generally, or in liver resident macrophages specifically, had no major effect on systemic glucose homeostasis and hepatic steatosis, inflammation, and fibrosis in this murine dietary model of MASLD/MASH. The complement system is a complex entity directing an important part of the body’s inflammatory and tissue repair response in MASLD. Further work is needed to elucidate the mechanisms of the role of C3aR1 in the pathogenesis of MASH and cirrhosis.

## Materials and Methods

### Animals

*C3ar1 flox/flox* mice were on the C57BL/6J background as described^43^. Homozygous LysM-Cre mice on the C57BL/6J background (Strain #004781) as well as homozygous Clec4f-Cre mice on the C57BL/6J background (Strain #003296) were purchased from Jackson Laboratories. *C3ar1 flox/flox* homozygous mice on C57BL/6J background were used in the experiments as controls from the same backcross generation^39^. All mice were maintained in plastic cages under a 12h/12h light/dark cycle at constant temperature (22°C) with free access to water and food. Mice were fed regular diet containing 4.5%kcal fat PicoLab Rodent diet 20 (LabDiet) or GAN diet containing 40%kcal HFD (mostly palm oil) with 20% fructose and 2% cholesterol (D09100310, Research Diets) for 28-30 weeks. Fat mass and lean mass were determined via noninvasive 3-in-1 body composition analyzer (EchoMRI). Mice were humanely euthanized with CO_2_ inhalation followed by exsanguination by cardiac puncture.

### Blood chemistry and serum insulin analysis

Mice were fasted overnight (14-16 hours) for glucose tolerance tests and injected intraperitoneally with syringe-filtered D-glucose solution (2g/kg). For insulin tolerance test, mice were fasted for 6 hours and injected with 0.5 mIU/kg insulin. Blood glucose levels were assayed by commercial glucometer (OneTouch) by tail vein blood samples. Plasma insulin levels were measured from mice fasted for 6 hours. Tail vein blood was collected into lithium heparin-coated tubes, centrifuged at 2000xg at 4°C, and plasma insulin levels were determined by ELISA using a standard curve (Mercodia). Serum alanine aminotransferase levels were measured in serum from blood collected via cardiac puncture using a commercially available colorimetric assay (TR71121, ThermoFisher Scientific).

### Peritoneal macrophage isolation and flow cytometry

Peritoneal macrophages were isolated from as previously described^44^. Briefly, mice were euthanized then immediately injected intraperitoneally with 10 mL phosphate-buffered saline (PBS, pH 7.4) at room temperature. After a 3-5 minute incubation period, peritoneal fluid was removed with sterile needle and syringe and placed on ice. After centrifugation at 300xg, the pellet was resuspended in PBS containing 2% fetal bovine serum and 0.1% sodium azide. Cells were stained with phycoerythrin-conjugated anti-F4/80 (clone BM8, cat. #123110) and fluorescein isothiocyanate-conjugated anti-CD11b (clone M1/70, cat. #101206) fluorescent antibodies (Biolegend). Stained cells were loaded on MA900 fluorescence-activated cell sorter (Sony), and dual-positive F480+/CD11b+ cells were sorted for subsequent RNA extraction.

### Histological studies

A mid-distal portion of the left liver lobe was fixed with 10% buffered formalin and transferred to 70% ethanol. Samples were embedded in paraffin, sectioned at ∼5mm thickness, and stained with Masson’s trichrome. Slides were imaged using Zeiss Axioscan7 at 20x magnification. Histologic analyses were performed using ImageJ software (version 1.53t). Lipid droplet area was quantified by subtracting non-droplet area in the green channel from total section area of 2-3 independent sections. Collagen proportionate area was quantified by measuring total area in the red channel after reducing intensity threshold to 60-70.

### RNA extraction and real-time quantitative PCR analysis

Total RNA from liver tissue lysates was extracted using Trizol reagent (Invitrogen) followed by RNAeasy Mini kit (Qiagen) as per manufacturer’s protocol. RNA was reverse-transcribed using the High Capacity cDNA RT kit (Thermo). Quantitative PCR was performed using SYBR Green Master Mix (Quanta) and specific gene primers on QuantStudio6 Flex Real-Time PCR Systems (Thermo Fisher Scientific) using the delta-delta Ct method. Expression levels were normalized to Ribosomal protein S18 (*Rps18*). Primer sequences are listed in Supplementary Table A.

### Statistical analyses

All statistical analyses were performed using GraphPad Prism10. Unpaired two-tailed Student’s *t* test with Welch correction for most analyses, with Holm-Šídák correction for multiple comparisons where applicable, and p<0.05 was considered statistically significant.

## Acknowledgments

We would like to thank Dr. Baran Ersoy, Dr. Robert Schwartz, and Dr. Saloni Sinha for their technical advice and assistance.

## Data Availability

Data will be made available upon reasonable request.

---

**S1).**
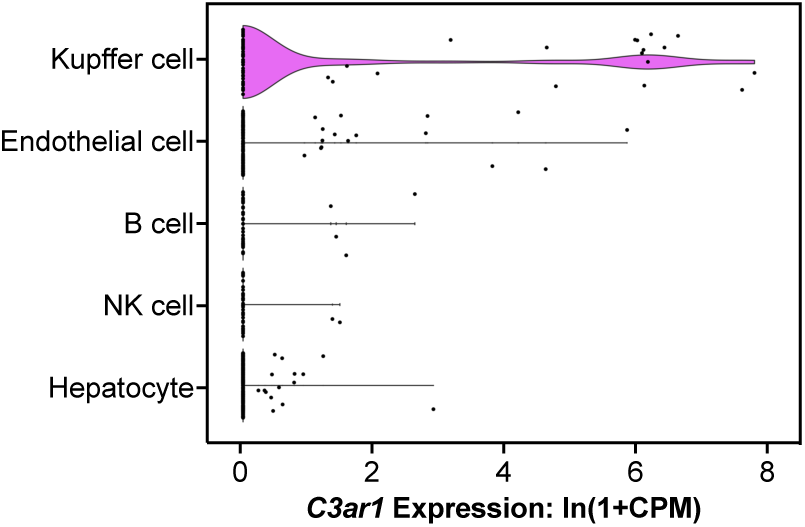
Single cell RNA sequencing analysis of *C3ar1* expression in mouse liver tissue (see text).

**S2).**
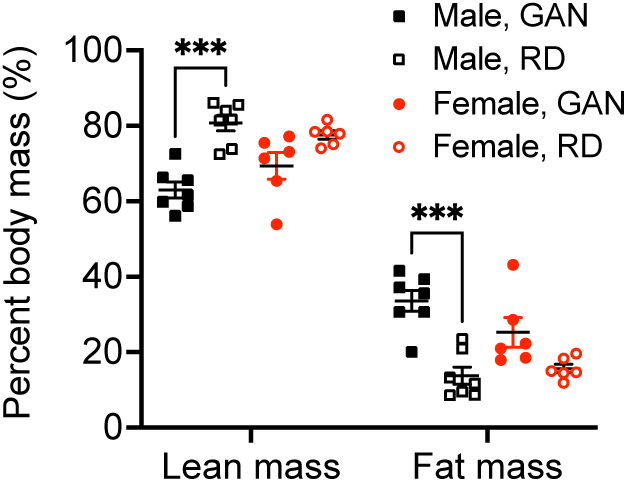
Percent lean and fat mass of flox/flox control mice after 20 weeks of GAN or RD (n = 6-7 per group).

**S3).**
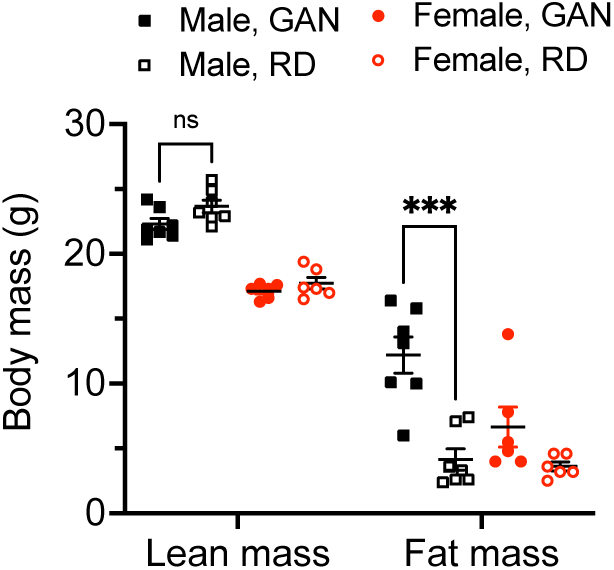
Absolute lean and fat mass of flox/flox control mice after 20 weeks of GAN or RD (n = 6-7 per group).

**S4).**
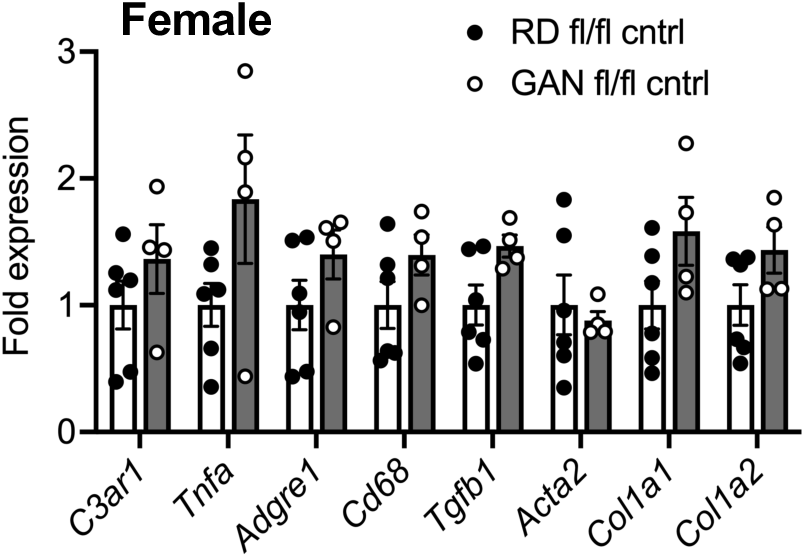
Relative gene expression in control female mice after 33 weeks on RD (n = 4-6 per group).

**S5).**
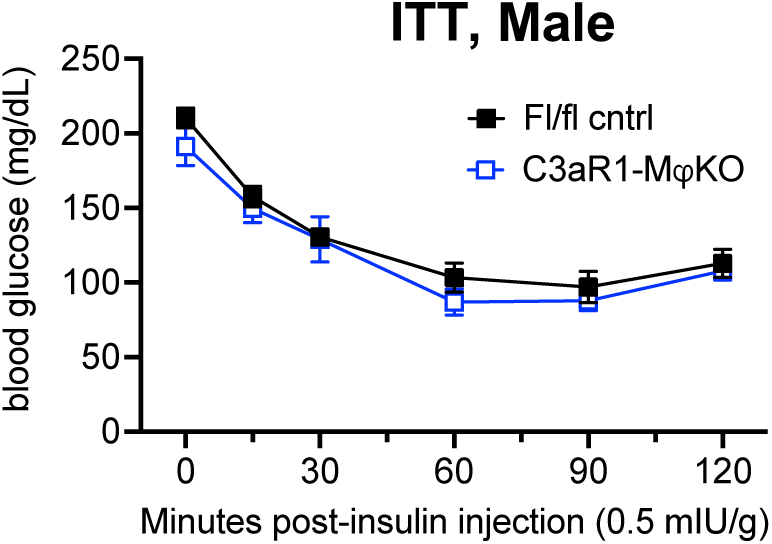
Insulin tolerance test in control or C3aR1-MjKO male mice with 14h fast after 29 weeks GAN diet (n = 6-9 per group).

**S6).**
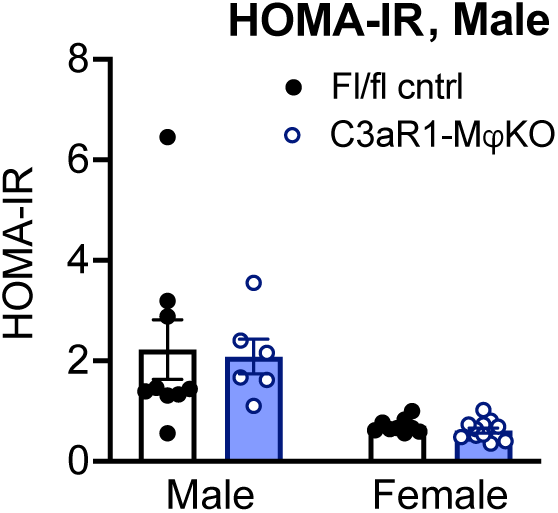
HOMA-IR measurement of insulin resistance in control or C3aR1-MjKO mice with 6h fast after 27 weeks GAN diet (n = 6-9 per male group, n = 9-13 per female group).

**S7).**
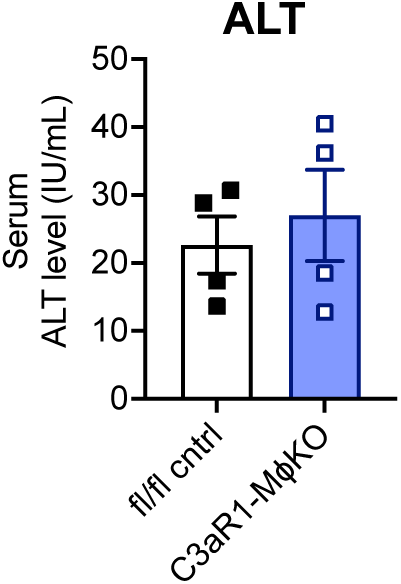
Serum alanine aminotransferase levels in control or C3aR1-MjKO male mice after 30 weeks GAN diet (n = 4 per group).

**S8).**
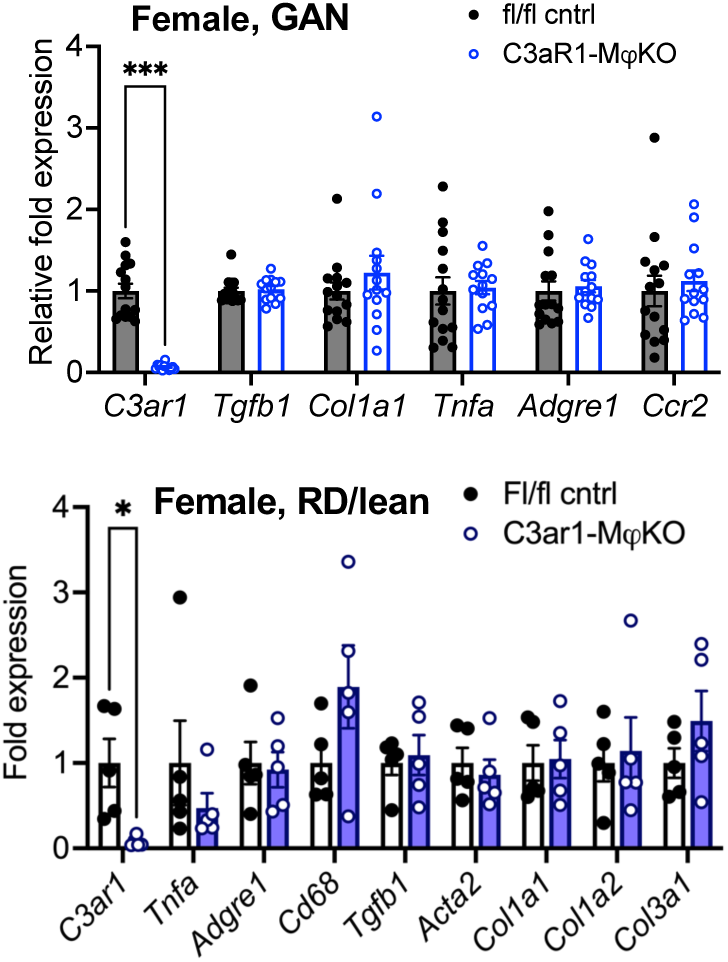
Relative gene expression in control or C3aR1-MjKO female mice after 30 weeks of either GAN (n = 13-14 per group) or RD (n = 5-6 per group).

**S9).**
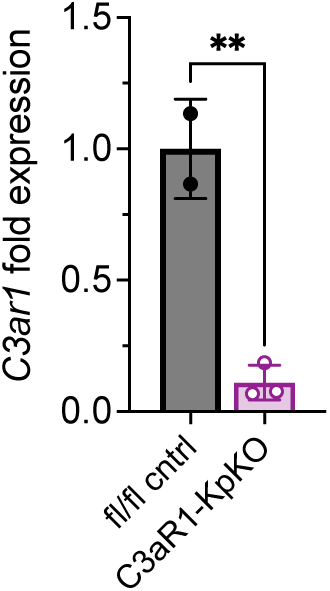
Relative *C3ar1* expression in control or C3aR1-KpKO female mice after 30 weeks RD (n = 2-3 per group). Unpaired two-tailed Student’s *t* test: **, p < 0.01; ***, p < 0.001.

**Supplementary Table S1.**
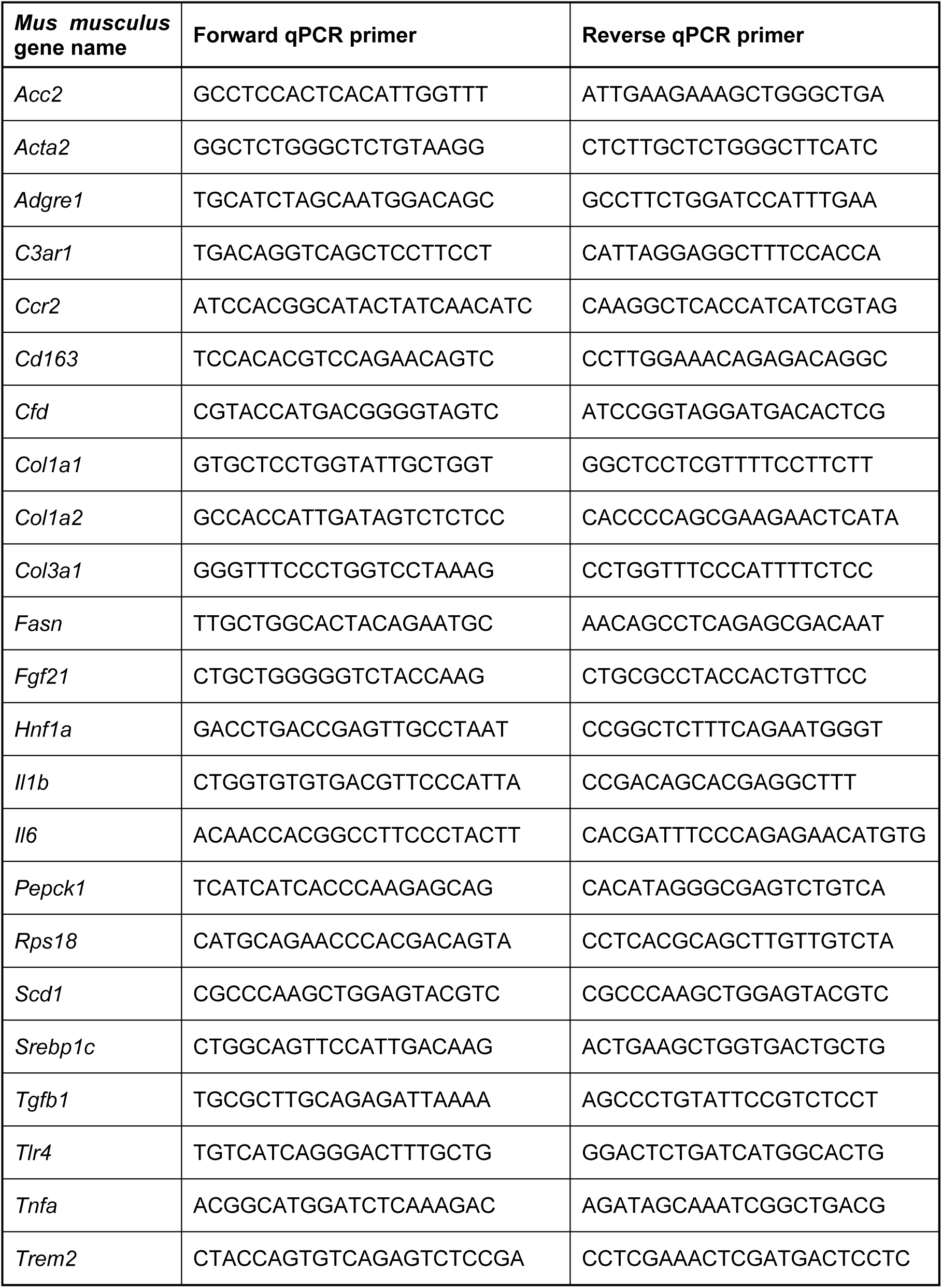

## References

1 Ge, X., Zheng, L., Wang, M., Du, Y. & Jiang, J. Prevalence trends in non-alcoholic fatty liver disease at the global, regional and national levels, 1990-2017: a population-based observational study. BMJ Open 10, e036663 (2020). 10.1136/bmjopen-2019-036663

2 Younossi, Z. et al. Global burden of NAFLD and NASH: trends, predictions, risk factors and prevention. Nat Rev Gastroenterol Hepatol 15, 11–20 (2018). 10.1038/nrgastro.2017.109

3 Ferguson, D. & Finck, B. N. Emerging therapeutic approaches for the treatment of NAFLD and type 2 diabetes mellitus. Nat Rev Endocrinol 17, 484–495 (2021). 10.1038/s41574-021-00507-z

4 Friedman, S. L., Neuschwander-Tetri, B. A., Rinella, M. & Sanyal, A. J. Mechanisms of NAFLD development and therapeutic strategies. Nat Med 24, 908–922 (2018). 10.1038/s41591-018-0104-9

5 Stefan, N., Haring, H. U. & Cusi, K. Non-alcoholic fatty liver disease: causes, diagnosis, cardiometabolic consequences, and treatment strategies. Lancet Diabetes Endocrinol 7, 313–324 (2019). 10.1016/S2213-8587(18)30154-2

6 Kim, H. et al. Metabolic Spectrum of Liver Failure in Type 2 Diabetes and Obesity: From NAFLD to NASH to HCC. Int J Mol Sci 22 (2021). 10.3390/ijms22094495

7 Duell, P. B. et al. Nonalcoholic Fatty Liver Disease and Cardiovascular Risk: A Scientific Statement From the American Heart Association. Arterioscler Thromb Vasc Biol 42, e168–e185 (2022). 10.1161/ATV.0000000000000153

8 Kasper, P. et al. NAFLD and cardiovascular diseases: a clinical review. Clin Res Cardiol 110, 921–937 (2021). 10.1007/s00392-020-01709-7

9 Barreby, E., Chen, P. & Aouadi, M. Macrophage functional diversity in NAFLD - more than inflammation. Nat Rev Endocrinol 18, 461–472 (2022). 10.1038/s41574-022-00675-6

10 Cai, J., Zhang, X. J. & Li, H. The Role of Innate Immune Cells in Nonalcoholic Steatohepatitis. Hepatology 70, 1026–1037 (2019). 10.1002/hep.30506

11 Guilliams, M. & Scott, C. L. Liver macrophages in health and disease. Immunity 55, 1515–1529 (2022). 10.1016/j.immuni.2022.08.002

12 Park, S. J., Garcia Diaz, J., Um, E. & Hahn, Y. S. Major roles of kupffer cells and macrophages in NAFLD development. Front Endocrinol (Lausanne) 14, 1150118 (2023). 10.3389/fendo.2023.1150118

13 Sakai, M. et al. Liver-Derived Signals Sequentially Reprogram Myeloid Enhancers to Initiate and Maintain Kupffer Cell Identity. Immunity 51, 655–670 e658 (2019). 10.1016/j.immuni.2019.09.002

14 Krenkel, O. et al. Myeloid cells in liver and bone marrow acquire a functionally distinct inflammatory phenotype during obesity-related steatohepatitis. Gut 69, 551–563 (2020). 10.1136/gutjnl-2019-318382

15 Guilliams, M. et al. Spatial proteogenomics reveals distinct and evolutionarily conserved hepatic macrophage niches. Cell 185, 379–396 e338 (2022). 10.1016/j.cell.2021.12.018

16 Govaere, O. et al. Transcriptomic profiling across the nonalcoholic fatty liver disease spectrum reveals gene signatures for steatohepatitis and fibrosis. Sci Transl Med 12 (2020). 10.1126/scitranslmed.aba4448

17 Merle, N. S., Church, S. E., Fremeaux-Bacchi, V. & Roumenina, L. T. Complement System Part I - Molecular Mechanisms of Activation and Regulation. Front Immunol 6, 262 (2015). 10.3389/fimmu.2015.00262

18 Kolev, M. & Kemper, C. Keeping It All Going-Complement Meets Metabolism. Front Immunol 8, 1 (2017). 10.3389/fimmu.2017.00001

19 Zhao, J. et al. Association of complement components with the risk and severity of NAFLD: A systematic review and meta-analysis. Front Immunol 13, 1054159 (2022). 10.3389/fimmu.2022.1054159

20 Segers, F. M. et al. Complement alternative pathway activation in human nonalcoholic steatohepatitis. PLoS One 9, e110053 (2014). 10.1371/journal.pone.0110053

21 Markiewski, M. M. & Lambris, J. D. The role of complement in inflammatory diseases from behind the scenes into the spotlight. Am J Pathol 171, 715–727 (2007). 10.2353/ajpath.2007.070166

22 Yadav, M. K. et al. Molecular basis of anaphylatoxin binding, activation, and signaling bias at complement receptors. Cell 186, 4956–4973 e4921 (2023). 10.1016/j.cell.2023.09.020

23 Flier, J. S., Cook, K. S., Usher, P. & Spiegelman, B. M. Severely impaired adipsin expression in genetic and acquired obesity. Science 237, 405–408 (1987). 10.1126/science.3299706

24 Xu, Y. et al. Complement activation in factor D-deficient mice. Proc Natl Acad Sci U S A 98, 14577–14582 (2001). 10.1073/pnas.261428398

25 Lim, J. et al. C5aR and C3aR antagonists each inhibit diet-induced obesity, metabolic dysfunction, and adipocyte and macrophage signaling. FASEB J 27, 822–831 (2013). 10.1096/fj.12-220582

26 Polyzos, S. A., Kountouras, J. & Mantzoros, C. S. Adipokines in nonalcoholic fatty liver disease. Metabolism 65, 1062–1079 (2016). 10.1016/j.metabol.2015.11.006

27 Han, J. & Zhang, X. Complement Component C3: A Novel Biomarker Participating in the Pathogenesis of Non-alcoholic Fatty Liver Disease. Front Med (Lausanne) 8, 653293 (2021). 10.3389/fmed.2021.653293

28 Lo, J. C. et al. Adipsin is an adipokine that improves beta cell function in diabetes. Cell 158, 41–53 (2014). 10.1016/j.cell.2014.06.005

29 Gomez-Banoy, N. et al. Adipsin preserves beta cells in diabetic mice and associates with protection from type 2 diabetes in humans. Nat Med 25, 1739–1747 (2019). 10.1038/s41591-019-0610-4

30 Mamane, Y. et al. The C3a anaphylatoxin receptor is a key mediator of insulin resistance and functions by modulating adipose tissue macrophage infiltration and activation. Diabetes 58, 2006–2017 (2009). 10.2337/db09-0323

31 Han, J. et al. Bone marrow-derived macrophage contributes to fibrosing steatohepatitis through activating hepatic stellate cells. J Pathol 248, 488–500 (2019). 10.1002/path.5275

32 Boland, M. L. et al. Towards a standard diet-induced and biopsy-confirmed mouse model of non-alcoholic steatohepatitis: Impact of dietary fat source. World J Gastroenterol 25, 4904–4920 (2019). 10.3748/wjg.v25.i33.4904

33 Hansen, H. H. et al. Human translatability of the GAN diet-induced obese mouse model of non-alcoholic steatohepatitis. BMC Gastroenterol 20, 210 (2020). 10.1186/s12876-020-01356-2

34 Vacca, M. et al. An unbiased ranking of murine dietary models based on their proximity to human metabolic dysfunction-associated steatotic liver disease (MASLD). Nat Metab (2024). 10.1038/s42255-024-01043-6

35 Uhlen, M. et al. Proteomics. Tissue-based map of the human proteome. Science 347, 1260419 (2015). 10.1126/science.1260419

36 MacParland, S. A. et al. Single cell RNA sequencing of human liver reveals distinct intrahepatic macrophage populations. Nat Commun 9, 4383 (2018). 10.1038/s41467-018-06318-7

37 Tabula Muris, C., et al. Single-cell transcriptomics of 20 mouse organs creates a Tabula Muris. Nature 562, 367–372 (2018). 10.1038/s41586-018-0590-4

38 Suppli, M. P. et al. Hepatic transcriptome signatures in patients with varying degrees of nonalcoholic fatty liver disease compared with healthy normal-weight individuals. Am J Physiol Gastrointest Liver Physiol 316, G462–G472 (2019). 10.1152/ajpgi.00358.2018

39 Ma, L. et al. Adipsin and Adipocyte-derived C3aR1 Regulate Thermogenic Fat in a Sex-dependent Fashion. JCI Insight (2024). 10.1172/jci.insight.178925

40 Kong, L. R. et al. Loss of C3a and C5a receptors promotes adipocyte browning and attenuates diet-induced obesity via activating inosine/A2aR pathway. Cell Rep 42, 112078 (2023). 10.1016/j.celrep.2023.112078

41 Li, X. et al. A new NASH model in aged mice with rapid progression of steatohepatitis and fibrosis. PLoS One 18, e0286257 (2023). 10.1371/journal.pone.0286257

42 Tsuchida, T. et al. A simple diet-and chemical-induced murine NASH model with rapid progression of steatohepatitis, fibrosis and liver cancer. J Hepatol 69, 385–395 (2018). 10.1016/j.jhep.2018.03.011

43 Cumpelik, A. et al. Dynamic regulation of B cell complement signaling is integral to germinal center responses. Nat Immunol 22, 757–768 (2021). 10.1038/s41590-021-00926-0

44 Zhang, X., Goncalves, R. & Mosser, D. M. The isolation and characterization of murine macrophages. Curr Protoc Immunol **Chapter** 14, 14 11 11–14 11 14 (2008). 10.1002/0471142735.im1401s83

